# A Transformer-Based Model Trained on Large Scale Claims Data for Prediction of Severe COVID-19 Disease Progression

**DOI:** 10.1101/2022.11.29.22282632

**Authors:** Manuel Lentzen, Thomas Linden, Sai Veeranki, Sumit Madan, Diether Kramer, Werner Leodolter, Holger Fröhlich, COPERIMOplus

## Abstract

In situations like the COVID-19 pandemic, healthcare systems are under enormous pressure as they can rapidly collapse under the burden of the crisis. Machine learning (ML) based risk models could lift the burden by identifying patients with high risk of severe disease progression. Electronic Health Records (EHRs) provide crucial sources of information to develop these models because they rely on routinely collected healthcare data. However, EHR data is challenging for training ML models because it contains irregularly timestamped diagnosis, prescription, and procedure codes. For such data, transformer-based models are promising. We extended the previously published Med-BERT model by including age, sex, medications, quantitative clinical measures, and state information. After pre-training on approximately 988 million EHRs from 3.5 million patients, we developed models to predict Acute Respiratory Manifestations (ARM) risk using the medical history of 80,211 COVID-19 patients. Compared to XGBoost and Random Forests, our transformer-based models more accurately forecast the risk of developing ARM after COVID-19 infection. We used Integrated Gradients and Bayesian networks to understand the link between the essential features of our model. Finally, we evaluated adapting our model to Austrian in-patient data. Our study highlights the promise of predictive transformer-based models for precision medicine.

## 1. Introduction

Coronavirus disease 2019 (COVID-19) is an infectious disease caused by severe acute respiratory syndrome coronavirus type 2 (SARS-CoV-2) that arose in December 2019. Since its emergence, 628 million people have been infected, and 6.58 million have died (https://coronavirus.jhu.edu/map.html, accessed 25.10.2022). In such pandemic circumstances, healthcare systems face a tremendous challenge as they can quickly collapse under the burden of this unprecedented crisis. Despite taking countermeasures such as testing, lockdowns, and vaccinations, the pandemic temporarily put immense stress on global healthcare systems. The use of decision support systems such as patient-level risk models can assist with the critical tasks of quickly and efficiently identifying high-risk patients so that the existing resources are best distributed and vulnerable patient subgroups are effectively protected.

Structured Electronic Health Records (EHRs) offer great opportunities for the efficient development of such risk models as they are routinely collected in many healthcare systems in large quantities. They contain data on diagnoses, prescriptions, procedures, and quantitative clinical measurements, such as vital values from bedside monitoring. Furthermore, demographic information such as age, gender, and region may be available. Models trained on such data could be used to better understand risk factors, such as comorbidities and medications, in addition to predicting a patient’s risk of severe disease development. However, it is difficult to exploit such data due to their high dimensionality, heterogeneity, temporal dependence, sparsity, and irregularity[1].

Furthermore, the coding of diagnoses is frequently biased for economic reasons. Since there is no unique mapping of a physician’s diagnosis to a coding scheme such as ICD, there is a tendency to select the code that delivers the greatest economic benefit from among several possible codes. Furthermore, medications are often categorized on a product level rather than a chemical substance level, and it is worth noting that several medications may contain the same chemical substance.

In the past, many ML approaches have been taken to work with structured EHR data. Simpler methods often limited the time information and just worked with a one-hot encoding (OHE) of diagnoses and prescriptions, which allowed the application of standard ML techniques, such as logistic regression, random forest (RF), XGBoost (XGB), and Bayesian methods[2]. Recently, more studies focused on the use of time-series information. Methods for such an approach include autoencoders, convolutional neural networks[3], or sequential models like recurrent neural networks (RNN)[4] or transformer-based models[5–9]. Transformer-based models originate from natural language processing (NLP) and have recently gained much attention since they have achieved excellent results in many areas[10–14]. A principal advantage of transformer models is the ability to train them in a parallel fashion and the ability to weigh different parts of a time series differently due to their inbuilt attention mechanism. Typically, transformer-based models are trained in two stages: a pre-training phase focusing on generic representation learning and a transfer-learning (fine-tuning) phase focusing on an application-specific prediction task. This approach has the advantage that pre-trained models, which are often trained on very large datasets (e.g., entire Wikipedia, all protein sequences), can be shared with a broader community and later on be fine-tuned for various tasks, which cannot be foreseen at the time of pre-training.

Variants of the Bidirectional Encoder Representations from Transformers (BERT)[15] model have recently been applied to structured EHR data. For instance, Shang et al. developed a graph-augmented transformer model named G-BERT to encode the medical history of single medical appointments and used the generated embeddings for a medication recommendation task[9]. Later, Li et al. developed BERT for EHR (BEHRT), which generated a patient embedding based on the history of diagnoses and used it for disease prediction in different time windows[5]. Since BEHRT – like most transformer-based models – is limited with respect to the maximum sequence length, the authors later developed a hierarchical BEHRT variant (HI-BEHRT), which can process longer medical histories[6]. Another model, called the Bidirectional Representation Learning model with a Transformer architecture on Multimodal EHR (BRLTM), was published by Meng et al. in 2021[8]. They followed a similar strategy as BEHRT but used a larger vocabulary, including more diagnoses, medications, and procedures. Another transformer-based model for structured EHR data is Med-BERT[7]. Like BRLTM, it is closely related to BEHRT, but Med-BERT has a much larger vocabulary size and uses slightly different training objectives. Unfortunately, none of the above-mentioned models is publicly available in a pre-trained form and thus not usable for the broader community.

Our contribution is an extension of the Med-BERT approach by including information about prescribed medications and demographic information such as state of residence, gender, and age as well as quantitative clinical measurements. We pre-trained our model, named ExMed-BERT, on 987,846,612 EHRs collected between 2010 and 2021, stemming from 3.5 million US patients in the IBM Explorys Therapeutic dataset. As a showcase, we subsequently used data from 80,211 COVID-19 patients to develop ML models for predicting the risk of acute respiratory manifestation (ARM) within three weeks after a confirmed COVID-19 diagnosis. This time frame was chosen because, on the one hand, a COVID-19 infection typically lasts 10 to 14 days. On the other hand, the timestamp of the COVID-19 diagnosis provided in the data may only be accurate up to a weekly resolution. The aim was thus to capture a serious event that could be time-wise related to the previously reported infection.

We compared our ExMed-BERT models with the two baseline models ignoring time information, RF and XGB. We then used explainable AI methods to gain insights into the underlying mechanisms of our models. A specific contribution is the use of Bayesian networks (BNs) to disentangle the relationship between most predictive features. Finally, we explored how our ExMed-BERT models could be adapted to external data from an Austrian hospital group (KAGes) via transfer learning strategies. Opposed to previous work, we make our ExMed-BERT model available to the scientific community.

## 2. Materials & Methods

### 2.1 General Overview

The work in this paper consists of four phases (Figure 1)

1. Pre-Training of transformer-based model for structured EHR data: Initially, we prepared a dataset of large-scale claims data and pre-trained a transformer-based model called ExMed-BERT for structured EHR data.
2. Development of risk models for COVID-19 disease progression: Subsequently, we used our newly trained model to develop risk models for predicting severe COVID-19 disease progression – namely ARM – and compared their performances with RF and XGB models.
3. Interpretation of developed risk models: Then, we used the Integrated Gradients approach in conjunction with Bayesian Networks to offer detailed explanations for model predictions.
4. Evaluation of the adaptation of our models to data obtained from an Austrian hospital group within a transfer learning approach.

**Figure 1.**
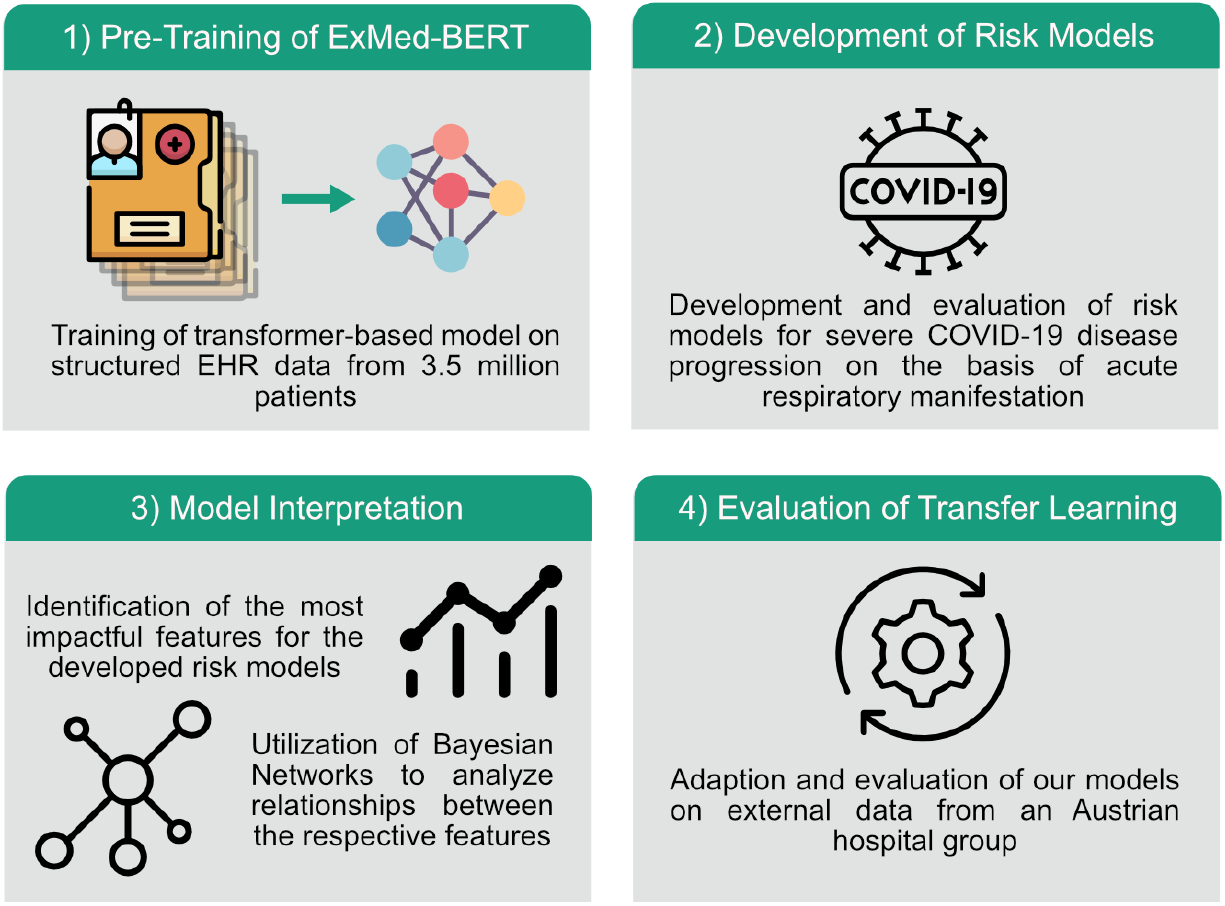
Study overview. First, we pre-trained a transformer model on about 988 million EHR records from 3.5 million patients. Then, we developed patient-level risk models for COVID-19 disease progression. Next, we interpreted our developed risk models using Integrated Gradients in conjunction with Bayesian networks. Finally, we evaluated the possibility of adapting the models to external data.

In the following, we describe our approach in more detail.

### 2.2 Data Preprocessing

#### 2.2.1 Preparation of Data for Modeling

This study used the IBM Explorys Therapeutic dataset (https://www.ibm.com/products/explorys-ehr-data-analysis-tools), which comprises EHRs and insurance claims from 4.5 million patients from all over the USA from 2010 until mid of 2021. Records consist of prescribed drugs, diagnoses, performed procedures, and a few quantitative clinical measures (e.g., blood pressure). We focused on demographic data and drugs, diagnoses, and available quantitative clinical measures. We excluded patients with fewer than five observations. This led to a reduced dataset of 3.5 million patients with 987,846,612 recorded diagnoses and drugs, which we used for pre-training a transformer model (details described later). The intent behind pre-training of a transformer model is to learn a suitable vector representation of time-stamped structured EHRs, irrespective of any later clinical use case. The fit of the model to a dedicated clinical endpoint is then performed within a subsequent fine-tuning / transfer learning step, for which we selected only patients with a confirmed COVID-19 diagnosis defined by the use of the International Classification of Diseases (ICD10)[16] code *U07*.*1* or a set of Logical Observation Identifier Names and Codes (LOINC)[17] codes (see Supplementary Section A) (n=80,211). We corrected the diagnosis or observation dates of the records by subtracting seven days to get an approximation of the index date of infection. Then we focused on the ARM endpoint, which was defined if at least one of the following diagnoses appeared within three weeks after the COVID-19 infection was reported (n=10,743):

- Pneumonia due to coronavirus disease 2019 (J12.82)
- Acute bronchitis due to other specified organisms (J20.8)
- Unspecified acute lower respiratory infection (J22)
- Bronchitis, not specified as acute or chronic (J40)
- Acute respiratory distress syndrome (J80)
- Respiratory failure, not elsewhere classified (J96)
- Other specified respiratory disorders (J98.8)

For fine-tuning, we used one year of medical history of the COVID-positive patients prior to their infection. Patients who fulfilled these criteria for the ARM endpoint were labeled as positives. Supplementary Figure A.1 depicts the filtering process in further detail.

To identify negatives while adjusting for the potentially confounding effects of age and gender, we used the technique of Inverse Probability of Treatment Weighting (IPTW)[18–20]. We used the Python package *psmpy*[21] (version 0.2.8) to calculate propensity scores (PS), and subsequently, the IPTW weights for each patient sample were calculated by the following equation and used in the fine-tuning process.

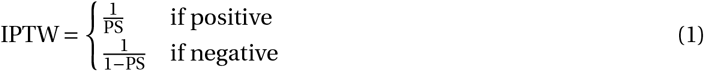

#### 2.2.2 Mapping of Drug and Diagnosis Codes

The IBM Explorys Therapeutic dataset includes information about diagnoses encoded as ICD9 and ICD10 codes and administered or prescribed drugs as RXNorm[22] identifiers. To harmonize the two versions of ICD diagnosis codes, we mapped them to Phecodes provided by the Phenome-wide association study (PheWAS)[23]. Due to the lower number of Phecodes, the problem of a non-unique mapping between a physician’s diagnosis and the ICD coding scheme is reduced. Hence, we reduced potential coding biases and reduced the feature space from 59,709 to 1,850 codes. Similarly, we mapped the provided RXNorm identifiers (RxCUI) to the fourth level of the Anatomical Therapeutic Chemical (ATC)[24] classification system for chemical compounds and thus addressed the sparse use of some RxCUIs by reducing the feature space from 23,801 to 630 codes.

#### 2.2.3 Input Representation for Pre-training

Unlike XGB and RF baseline models, our transformers require a pre-training phase. As transformer models are specifically designed to work on data of sequential nature, we need to represent the entire medical history of each patient as a sequence. We generated separate sequences for each modality, as depicted in the lower part of Figure 2. Each element of the sequence is an integer corresponding to one vector of the embedding matrices. Quantitative clinical measures were not considered at this point but only during the subsequent fine-tuning phase.

**Figure 2.**
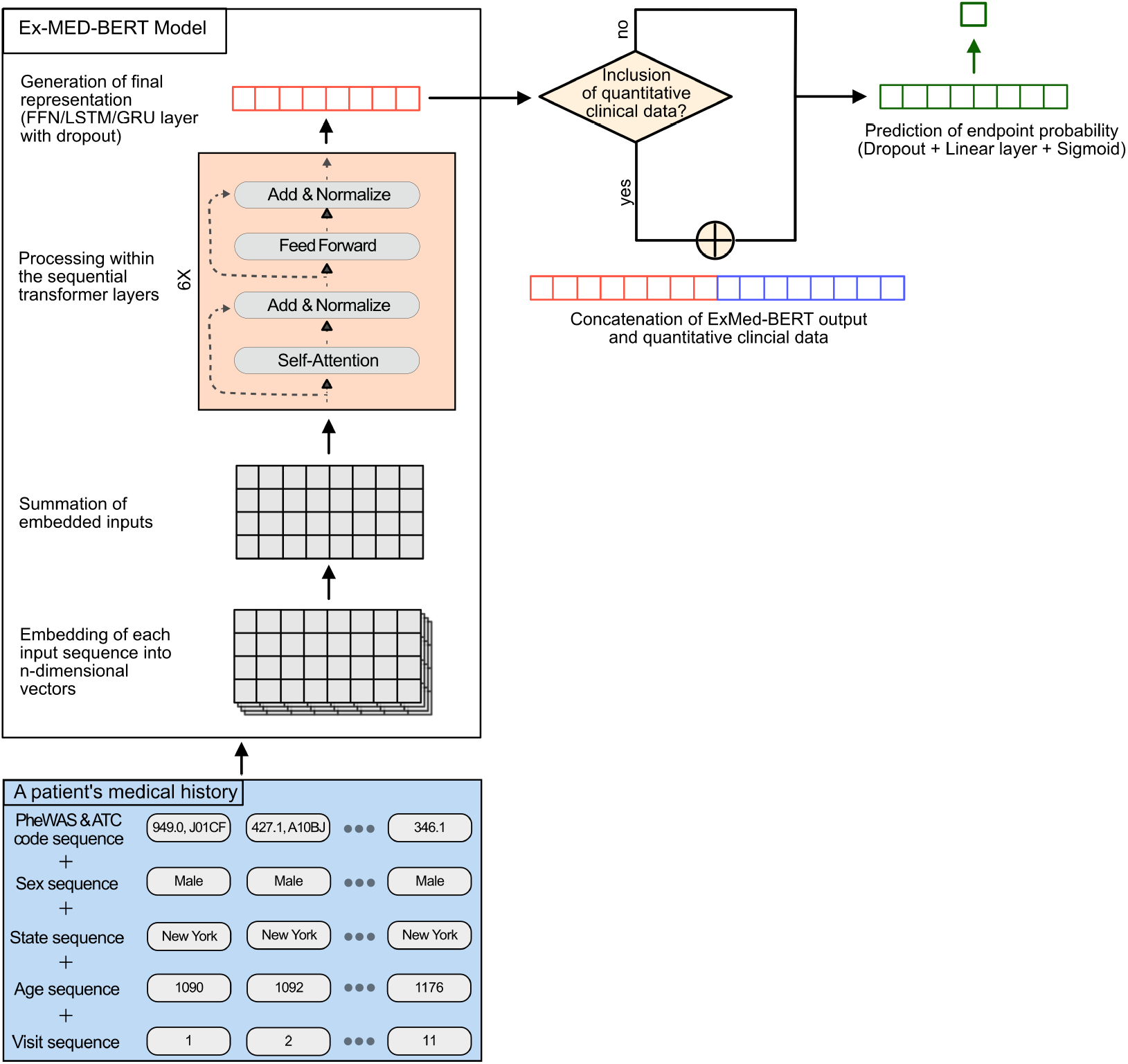
Overview of the model structure. Compared to BERT or other transformer-based models, we employed a multimodal embedding layer for structured EHR data comprising of drug, diagnosis, visit information, and information about a patient’s sex, state of residence, and age. After embedding, the input is passed through 6 transformer layers before a final representation of a patient’s medical history is generated with an FFN, LSTM, or GRU head. Subsequently, these patient representations were either concatenated with the quantitative clinical data or directly passed through an FFN head for classification.

### 2.3 Model Training and Evaluation

#### 2.3.1 Basic Model Structure and Pre-training of ExMed-BERT

We followed a similar strategy as the Med-BERT paper and focused on an extension of the BERT embedding layer. In addition to the diagnoses that were included in the Med-BERT model, we further extended Med-BERT by adding information on prescribed drugs, the patient’s sex, state of residency, and age. We denote our model as Extended Med-BERT (ExMed-BERT). As shown in Figure 2, we used different embeddings to accommodate the five feature modalities. Diagnoses and drugs were represented in one embedding via Phecodes and ATC codes. The sex and state embeddings contained static information. The age sequence contained the patients’ age encoded in months, and lastly, the visit sequence was used to distinguish between each visit in a sequence. Since the order of drugs and diagnoses within one visit was random, we passed on a serialization embedding. Similar to Med-BERT, we also did not use CLS and SEP tokens in our input sequences.

We used the same hyperparameters and training objectives as Med-BERT and pre-trained the model on the entire information of the 3.5 million patients in the pre-training cohort. If sequences exceeded the maximum sequence length of 512 diagnosis and drug codes, we split the sequences and processed the samples individually. We used the following joint training objectives to pre-train our model:

- **Masked language modeling (MLM)**: This task is identical to the BERT approach and we followed the Med-BERT strategy in masking only one of the codes at a time. In 80 % of the cases, the masked code was replaced with [MASK], in 10 % it was replaced with another code and in the remaining 10 %, it remained unchanged. The model’s task was to predict the correct code based on the information provided by the remaining sequence.
- **Prediction of prolonged length of stay (PLOS) in hospital**: As Rasmy et al.[7], we also predicted whether a patient had a prolonged stay in a hospital (>7 days) throughout his or her medical history. This task requires assessing the severity of a patient’s health condition throughout their medical history.

#### 2.3.2 Machine Learning-Based Risk Models

We developed ML-based risk models for the above-defined endpoint ARM using RF, XGB, and our ExMed-BERT architecture while at the same time adjusting for the potentially confounding effects of age and gender via the IPTW approach described before. During the fine-tuning of our ExMed-BERT model, we evaluated different classification head variants. We trained three models by using a feed-forward network (FFN), long short-term memory (LSTM), and gated recurrent unit (GRU) head to classify whether a patient was positive for the respective endpoint or not. We split the data into training, validation, and testing sets in a stratified manner (70/10/20 %) and used Bayesian hyperparameter optimization (*optuna* [25], version 2.10.0) to tune model parameters such as the learning rate, batch size, warmup ratio, weight decay, and in the case of RNNs, also the number of RNN layers. Similarly, we trained RF and XGB classifiers and optimized several model hyperparameters. A detailed list of all optimized parameters can be found in Supplementary Table B.1.

In the case of the RF and XGB models, we represented all categorical features in a one-hot encoding, meaning that if a diagnosis or a drug was recorded at any time point in the considered one-year medical history, it was labeled with one, if not it was labeled with zero. Similarly, we encoded the state of residence and sex.

#### 2.3.3 Combination with Quantitative Clinical Measurements

In this work, we also evaluated the combination of diagnosis and prescription codes with quantitative clinical data, such as blood pressure measurements. We restricted ourselves to data recorded in the two weeks prior to the corrected index date and excluded features with a missingness of more than 60 %. Due to the high sparsity of this data, we ended up with only eight features for our experiments: weight, body mass index (BMI), body surface area (BSA), body height, temperature, diastolic and systolic blood pressure, and heart rate. The number of patients with available quantitative data for each of these features is shown in Table 1 (n=23,949). We used an RF-based approach to impute the quantitative data for all patients while only using the training data (*missingpy*[26], version 0.2.0).

**Table 1.** Overview of the number of patients in the two datasets used in this study. ARM stands for acute respiratory manifestation.

|  | IBM Explorys<br>Therapeutic<br>Dataset | Data from an<br>Austrian Hospital<br>Group |
| --- | --- | --- |
| Entire dataset (unfiltered) | 4,563,769 | – |
| Pre-training cohort | 3,478,438 | – |
| Fine-tuning cohort | 80,211 | 6,335 |
| ARM patients | 10,743 | 385 |
| Patients with quantitative clinical data | 23,949 | – |

For the RF experiments, the quantitative clinical features were directly concatenated with the OHE. For the XGB model, we performed no prior imputation; instead, we directly concatenated the quantitative clinical features with the OHE and relied on the implicit imputation mechanism implemented in XGB.

While no changes were necessary for the RF and XGB models, we had to modify the ExMed-BERTs model architecture to handle quantitative clinical features. More specifically, we used the same ExMed-BERT model as before to generate embeddings of a patient’s medical history and concatenated those vector-based representations with the quantitative input before passing it into a final classification head (see Figure 2).

### 2.4 Model Interpretation

#### 2.4.1 Feature Importance

To better understand the ExMed-BERT models, we used the Integrated Gradients (IG)[27] approach to determine which drugs and diagnoses had the highest influence on the model predictions. The IG method is an axiomatic model interpretability technique that awards, in the case of the ExMed-BERT models, an attribution score for each diagnosis or drug in the medical history. Next to an input sample (*x ∈ Rn*), the IG method requires a baseline input (*x′ ∈ Rn*), which we constructed using a sequence of padding tokens. The IGs are then approximated by summing the gradients at points along the path from the specified baseline to the input using the following formula:

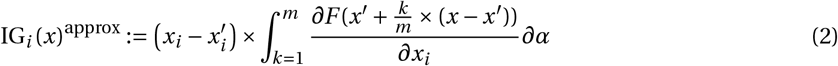

Here, *m* is the number of steps, and *F* is a function (*F* : *R*^*n*^ *⇒* [0, 1]) which represents our ExMed-BERT model. We performed 50 steps to approximate the integrated gradients.

Initially, we computed IG attributions for all patients in the test dataset. Based on these, we calculated the mean absolute attribution for each diagnosis and drug that occurred at least ten percent of the time to identify the top features for each model. Subsequently, we calculated partial dependency scores using the top 20 features. To do so, we first calculated the probability for each patient for a specific endpoint using our fine-tuned ExMed-BERT models; we refer to this probability as *pr*. The data for each of the top 20 features were then permuted individually by exchanging the respective diagnosis or drug codes with a *PAD* token. Subsequently, the modified data was used as input for our models to calculate the probability *pm*. Finally, a fold change for each feature was calculated using the probabilities obtained for actual (*pr*) and modified data (*pm*) to estimate the effect (fold change; FC) of certain features on the model’s prediction:

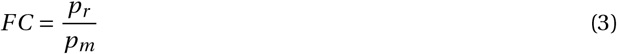

#### 2.4.3 Unraveling Feature Dependencies

To better comprehend the numerous interactions and dependencies between the most influential features, we developed BN models. BNs are probabilistic graphical models that can represent complex multivariate distributions with many variables. They can be graphically depicted with nodes representing random variables and edges expressing conditional statistical relationships. Let *G* = (*V,E*) be a directed acyclic graph and {*X*_*v*_ | ν ∈ *V*} a set of random variables indexed over nodes in V. Then for any *B* = (*X,G*):

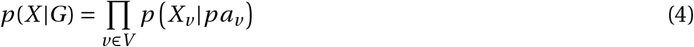

where *pa*_*v*_ denotes the parents of *v ∈ V* according to the graph structure *G*. Because of their ability to model (potentially causal) relationships between variables, BNs are frequently employed in many areas of science, including system biology and medicine. In this work, we learned the graph structure G of a BN for the 100 most important features (according to the IG method) using the R package *bnlearn*[28] (version 4.7). We used a one-hot encoding for the respective features to indicate whether it was present in the one-year medical history, similar to the data preparation for the tree-based models. We also provided the patients’ age, sex, and endpoint status. The tabu algorithm[29, 30] was used for BN structure learning. This was performed within a non-parametric bootstrap sampling scheme: We randomly subsampled n = 80,211 patients with replacement for 1000 times, and for each bootstrap sample we performed a complete network structure learning. We then focused on edges occurring in over half of the 1000 network architectures acquired from the non-parametric bootstrapped samples.

### 2.5 Transfer Learning on Austrian Hospital Data

#### 2.5.1 Overview about Data

Data from the Austrian hospital group consisted of pseudonymized in-patient records of 6,335 COVID-19 positive patients, out of which 385 suffered from ARM within a 3-week follow-up period after the initial visit to the hospital. The medication prescriptions were already encoded in ATC, but as ICD9/10 codes were used for diagnoses, these were mapped to Phecodes, akin to the procedure described earlier for the IBM Explorys dataset.

#### 2.5.2 Transfer Learning of ExMed-BERT

We continued training of the ExMed-BERT model for the ARM endpoint for only five epochs on the Austrian hospital data. This was done due to computational constraints. For the same reason, we did no substantial hyperparameter tuning but used the optimal hyperparameters discovered on the IBM Explorys data. We used 5-fold cross-validation to account for the small amount of available data. Alongside the ExMed-BERT model, we trained a new RF model as a comparison.

## 3. Results

In this study, we predicted severe COVID-19 disease progression based on a patient’s medical history. We begin by presenting the pre-training results of our newly created ExMed-BERT model. Then, we show the performances of the developed risk models, and lastly, we interpret our models using an explainable AI methodology.

## 3.1 Model Pre-Training

We utilized MLM and PLOS as training objectives for pre-training of the ExMed-BERT model. After 4.5M steps (epoch 37), the MLM accuracy increased to around 51 % and the PLOS F1-score to 70 %. Following the inclusion of 61 missing ATC codes and the corresponding changes to the embedding, we began training for 750K steps. Finally, we achieved an MLM accuracy of 67 % and a PLOS F1-score of 66 % (epoch 42, Supplementary Figure B.1).

### 3.2 Evaluation of Risk Models

Following pre-training, we developed and evaluated risk models for the prediction of the ARM endpoint. Initially, we considered only the medical history without additional quantitative clinical measures. As shown in Table 2, all ExMed-BERT models performed better than the RF and XGB variants on unseen test data. Without quantitative clinical data, the ExMed-BERT models scored roughly 78 % AUROC for the ARM endpoint, and the AUPR varied between 36.7 % and 38.2 %. The RF model, on the other hand, only achieved an AUROC of 73.4 % and an AUPR of 29.1 %. The XGB model had a slightly lower AUROC of 72.4 % and AUPR of 28.2 %.

**Table 2.**
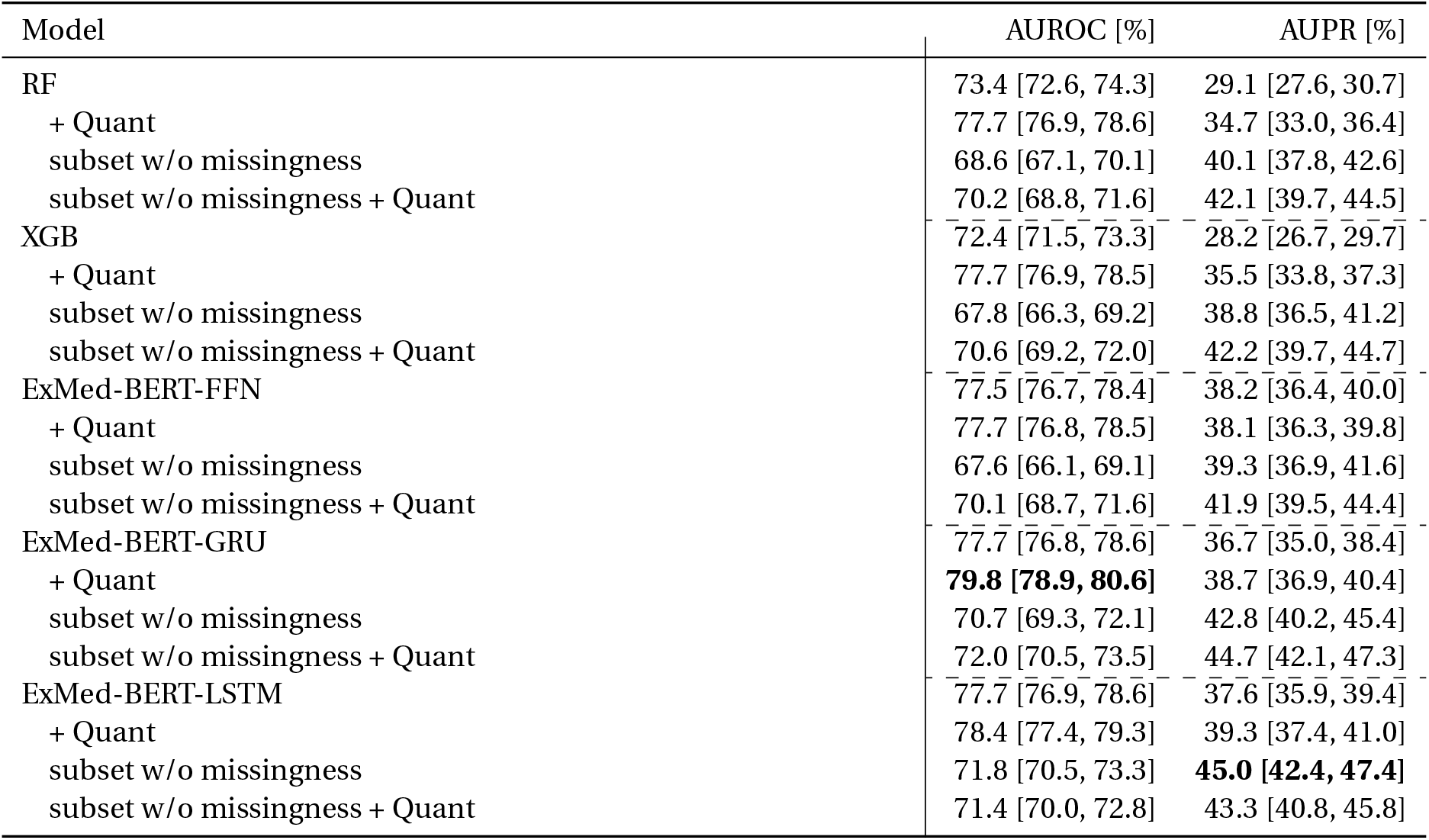
Evaluation results of the risk models for predicting ARM. Areas under the Receiver-Operator Characteristic Curve (AUROC) and the Precision-Recall Curve (AUPR) are shown for each model and feature modality. The best results per column are highlighted in bold. The suffix “+Quant” stands for the additional use of quantitative clinical data during fine-tuning. The suffix “subset w/o missingness” indicates that we used a reduced subset (n = 23,949) for training and evaluation, where all quantitative clinical measures were available. The values in brackets indicate the 95 % confidence interval which we estimated by performing bootstrap resampling for 1000 times.

The results of nearly all models improved when quantitative clinical measurements were integrated. The ExMed-BERT model with the GRU classification head integrating quantitative data gave the overall best result, with an AUROC of 79.8 % and an AUPR of 38.7 %, which is significantly higher than all other models.

When only patients with fully recorded quantitative clinical measurements were used, all models performed worse. That means the potential negative effect of imputing missing values was far less than the benefit of including additional data.

## 3.3 Model Explanation

To better understand model predictions, we used an explainable AI methodology – namely IG – to calculate attribution scores for all features in the best-performing ExMed-BERT model. We calculated the IG attributions and used them to identify the 20 most important features by ranking them based on their mean absolute value. Figure 3 shows all the IG attributions and FC scores, which are in agreement with each other. We found that the presence of diagnoses for chronic airway obstruction, congestive heart failure, cough, dementia, edema, obesity, shortness of breath, spondylosis, and type 2 diabetes in the medical history has a large impact on the prediction of a patient’s risk for ARM. Similarly, the prescriptions of angiotensin II receptor blockers, biguanides, dihydropyridine derivatives, and thiazides have a substantial positive impact on our models’ predictions.

**Figure 3.**
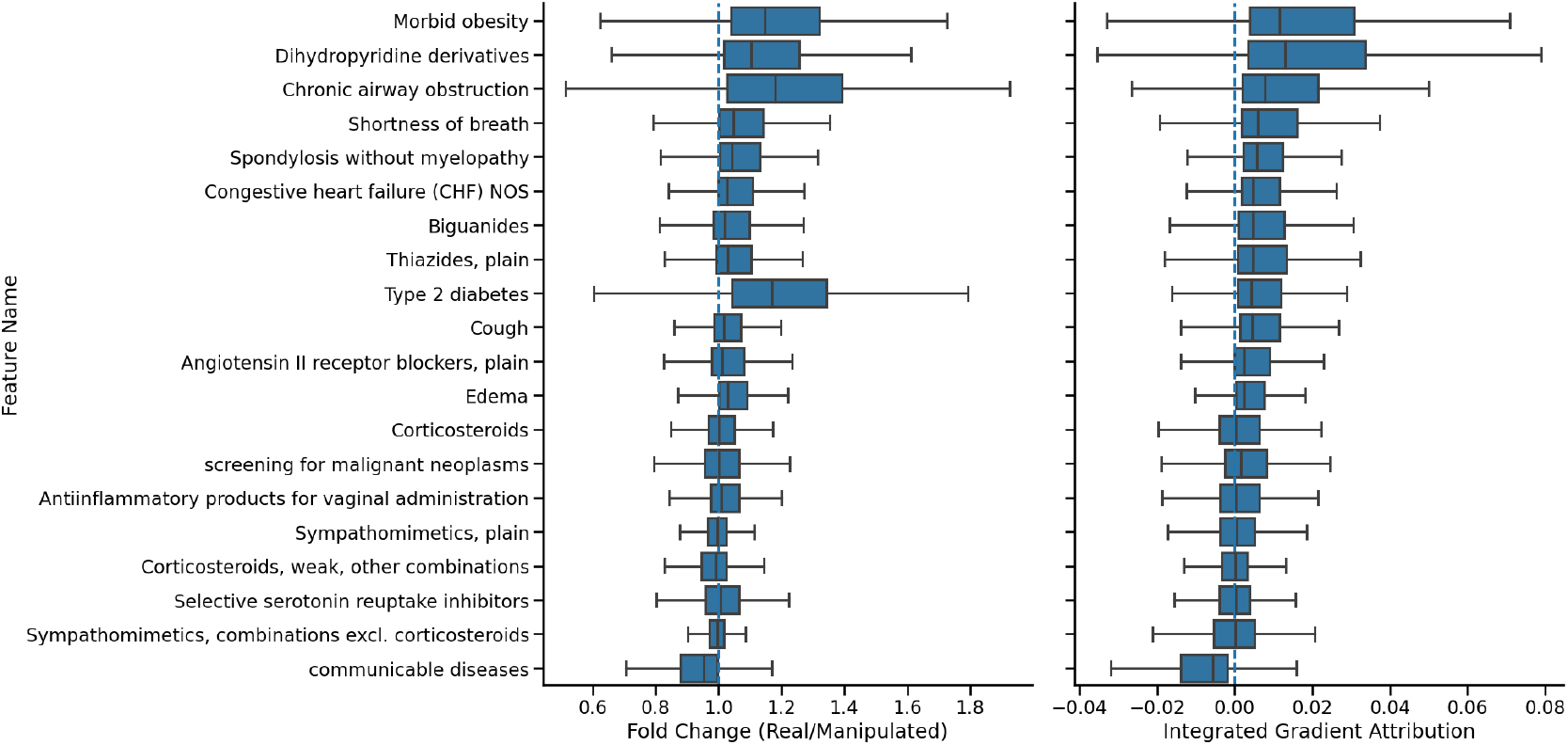
Integrated Gradients Attributions for ExMed-BERT GRU. Depicted are all calculated fold changes (FC) and IG attributions for the 20 most important features for the prediction of ARM onset. The dashed blue lines indicate neutral attributions. Everything greater than the neutral value has a positive effect on the prediction and vice versa.

Of course, these prescriptions and diagnoses could be correlated with each other, and thus, not all of them might have a direct impact on the ARM endpoint. Hence, we learned the graph structure of a BN to determine how the significant diagnoses or drugs could be related to one another. The overall network structure is provided as a *graphml* file, an XML-based data format for graph representation, as supplementary material to this paper. Figures 4 and 5 show two excerpts of the BN graph structure. Figure 4 focuses on Angiotensin II receptor blockers and their relationship to other drugs and diagnoses. Angiotensin II receptor blockers are used to treat hypertension, kidney diseases, and heart failure[31]. Furthermore, our graph shows a connection to essential hypertension and several ATC subgroups, namely ACE inhibitors, Dihydropyridine derivates, HMG CoA reductase inhibitors, and Thiazides.

**Figure 4.**
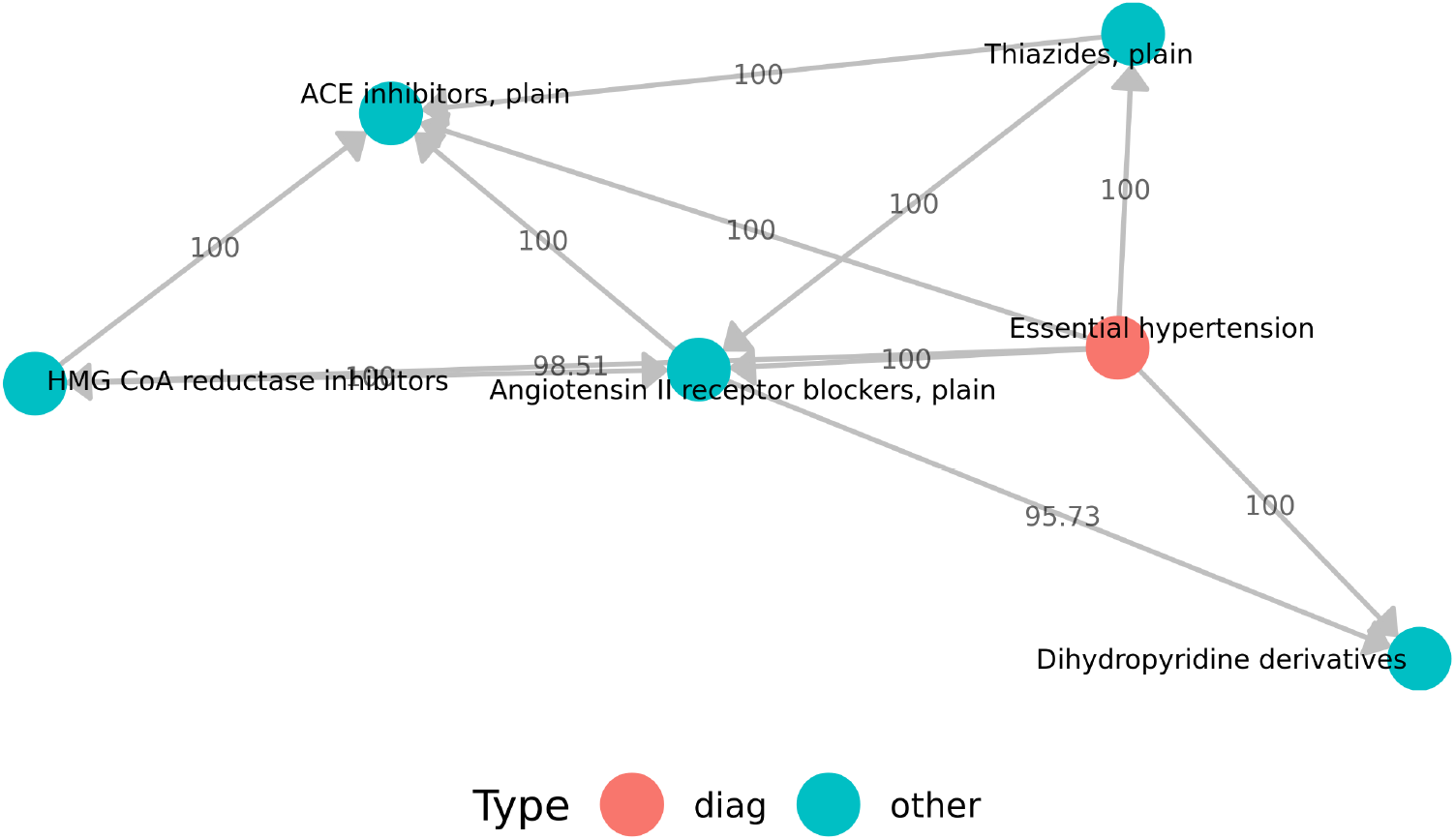
Angiotensin II receptor blockers and the direct neighborhood in the inferred Bayesian network. The numbers indicate the bootstrap strength of the respective edges in percentage. That means a bootstrap strength of 100 indicates that the corresponding edge has been found in each of 1000 BN reconstructions learned from different bootstrap samples.

**Figure 5.**
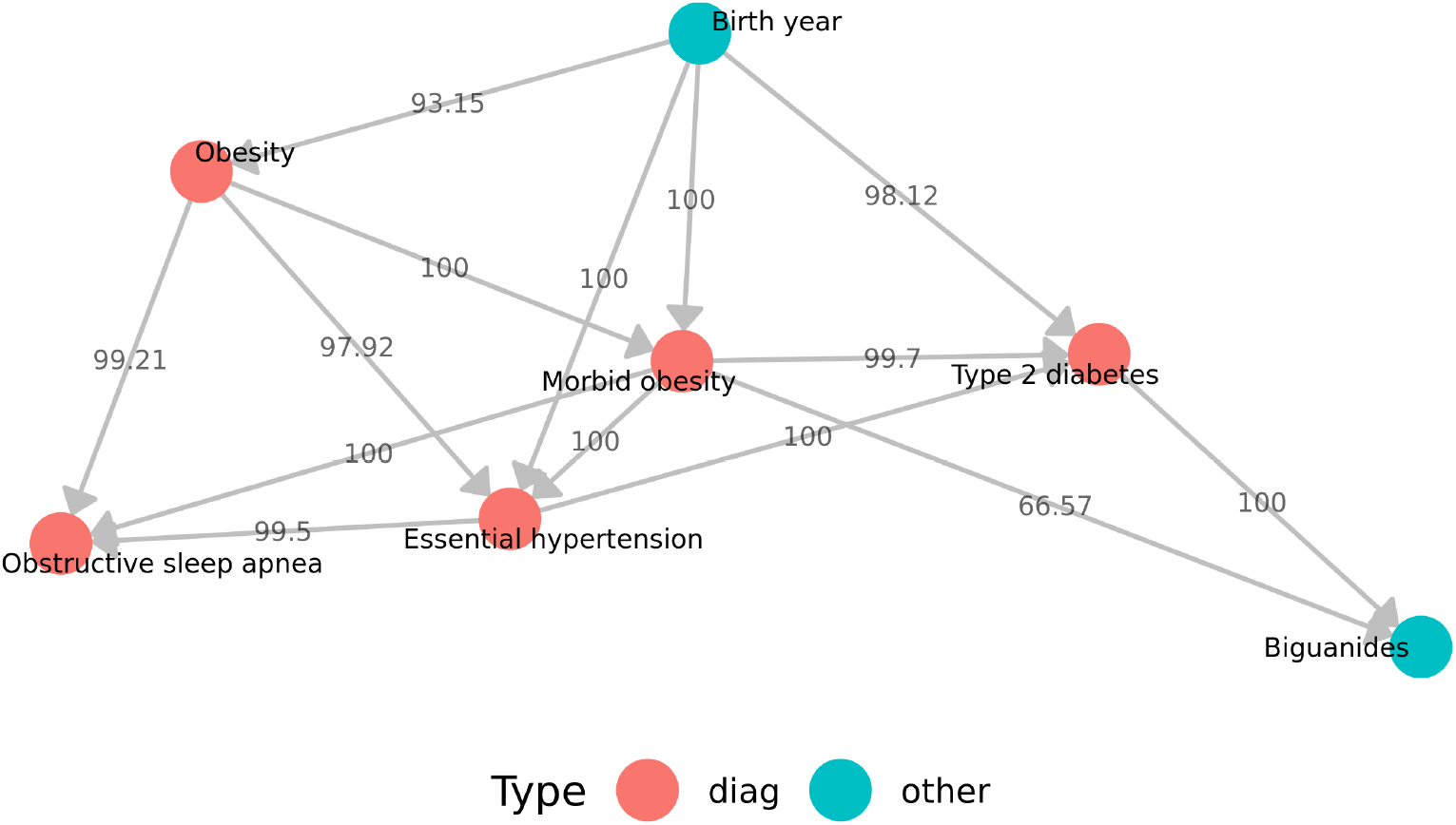
Morbid obesity and the direct neighborhood in the inferred Bayesian network. The numbers indicate the bootstrap strength of the respective edges in percentage. That means a bootstrap strength of 100 indicates that the corresponding edge has been found in each of 1000 BN reconstructions learned from different bootstrap samples.

Figure 5 depicts morbid obesity and other diagnoses and drugs in its immediate neighborhood. There is a link to the class of Biguanides, which includes the drug Metformin which is commonly used to treat diabetes[32]. Furthermore, morbid obesity is linked to hypertension, type 2 diabetes, obstructive sleep apnea, and obesity.

We aimed for an understanding of the statistical and potentially causal effect of those features on the endpoint, which were either among the 20 most important features or sink nodes in the BN. The latter are nodes without outgoing connections and, therefore, do not influence any other features according to our BN analysis. For each of those features, we performed a univariate logistic regression analysis while using IPTW case weights to correct for potential confounding effects of age and gender. Our analysis shows significant effects of several prior diagnoses on the ARM onset, namely, type 2 diabetes, obesity, dementia, cardiovascular diseases, and respiratory diseases (see Table 3). These morbidities have previously been reported as risk factors for severe COVID-19 disease progression[33–38], and also the underlying molecular mechanisms have been discussed[39, 40]. Besides, we found significant effects between constipation, screening for malignant neoplasms, and infectious/parasitic diseases and the ARM onset. This might be explained by the fact that such procedures are more frequently executed in older patients with a bad health condition, hence resulting in a higher risk of severe COVID-19 progression. In the same type of patients constipation is also a frequent problem, e.g., due to lifestyle.

**Table 3.** Features with a significant statistical effect on ARM. We performed a logistic regression and corrected for the confounding variables age, sex, and state of residency. The p-values were corrected for multiple testing using the Holm-Sidak method.

| Feature Names | Corrected p-value |
| --- | --- |
| Type 2 diabetes | <0.0001 |
| Heart failure with reduced EF | <0.0001 |
| Shortness of breath | <0.0001 |
| Constipation | <0.0001 |
| Morbid obesity | <0.0001 |
| Screening for malignant neoplasms | <0.0001 |
| Dementias | <0.0001 |
| Chronic airway obstruction | 0,0003 |
| Cough | 0,0018 |
| Screening for infectious and parasitic diseases | 0,0083 |
| Congestive heart failure (CHF) NOS | 0,0119 |

### 3.4 Transfer Learning on Austrian Hospital Data

Fine-tuning of Ex-MedBERT on a small set of pseudonymized in-patient data from an Austrian hospital group resulted in a prediction performance almost identical to the one observed for an RF trained de novo on the same data (Supplementary Table B.2). At the same time, prediction performances were significantly lower than the ones observed on IBM Explorys (AUC *≈* 60 %). We will elaborate on potential reasons in the subsequent discussion.

## 4. Discussion

Pandemics such as COVID-19 pose immense challenges to global healthcare systems. Utilizing patient-level risk models to support doctors and clinics is one way to maximize the use of available resources. Following previous research, we trained a transformer-based model on structured EHR data in this study. In contrast to the prior approaches, such as BEHRT or Med-BERT, we incorporated additional data modalities and developed risk models for COVID-19 disease progression. Prediction performances achieved by our ExMed-BERT model are altogether superior to those reported by Lazzarini et al. [41] for the closely related endpoint acute respiratory distress syndrome (ARDS)[41]. The authors trained an XGB based on US administrative claims data from 290,000 patients and achieved an AUROC of 69 % and an AUPR of 7 %. For comparison, using data from intensive care units (ICUs), Bendavid et al. reported an AUROC of 83 % for an XGB trained to predict the initiation of invasive mechanical ventilation[42], and Singhal et al. achieved an AUROC of 89 % for predicting the onset of ARDS[43]. Importantly, ICU data are structurally and content-wise very different from the data used in our study, which comprises in-patient as well as out-patient information over a longer period (here: one year), but only contains limited quantitative information. Altogether, our findings are in line with previous studies [5, 7, 8], showing that transformer-based models are well-suited for structured EHR data similar to ours. Even without additional quantitative information, our ExMed-BERT outperformed the RF and XGB models. With the inclusion of quantitative clinical measures, our ExMed-BERT models further increased in prediction performance. For that purpose, we proposed a novel approach to combine quantitative clinical measures with the embeddings of EHR codes learned by ExMed-BERT, which resulted in the overall best-performing model. Our results thus demonstrate the importance of combining diagnosis and prescription codes with quantitative clinical measures for developing risk models. Even though these quantitative clinical measures were only taken at a single time point in the two weeks preceding COVID-19 infection and not for every patient, using these data could provide better performance.

Using a combined strategy consisting of feature importance analysis, BN structure learning and statistical hypothesis testing, we were able to identify diagnoses and prescriptions that have a significant impact on model prediction and may causally influence the endpoint. Our analysis supports that socioeconomic and psycho-social health risks play an important role in addition to well-known risk factors such as obesity, diabetes, cardiovascular diseases, and dementia, which have already been reported as known risk factors for severe COVID-19 disease progression in several studies [33, 36, 44–46]. This confirms the validity of our approach, which can be applied to other datasets as well.

Our work demonstrates the potential of a transformer-based pre-training / fine-tuning strategy to develop risk models for precision medicine. This strategy provides the chance to perform transfer learning of our model on data from other organizations and thus use the pre-trained ExMed-BERT as a basis for future model development. Our experiment with data from an Austrian hospital group demonstrated the potential as well as the limitations of such an approach: The data from the Austrian hospital group only comprises in-patient information, and the number of patients is far smaller than during the fine-tuning phase on the IBM Explorys data (6,335 patients instead of 80,211). Furthermore, the ratio of ARM-positive patients is significantly lower (6.1 % instead of 13.4 %). Notably, there could also be different medical coding practices in the two countries. Finally, constraints on the technical equipment within the Austrian hospital group only allowed us to fine-tune our model for a small number of epochs and without hyperparameter tuning. Due to all these factors, our ExMed-BERT model fine-tuned on the Austrian data achieved a performance that was comparable to an RF model trained de novo on the same data but significantly lower than prediction performances achieved on US data. We thus conclude that having a sufficiently large dataset with a number of patients in a range comparable to the IBM Explorys data would be a prerequisite to obtaining better models in a transfer learning setting. Furthermore, appropriate technical equipment is important. Finally, integration of in-patient and out-patient data is required, at least for our model.

## 5. Conclusion

Our work demonstrates the potential of customized transformer-based models for analyzing structured EHR data. We showed that it is possible to integrate quantitative clinical data into such models, which can significantly improve prediction performance. Furthermore, we introduced a general approach for explaining ExMed-BERT model predictions. Transfer learning strategies open the possibility of leveraging our pretrained ExMed-BERT model for the prediction of clinical endpoints different from the one addressed within this paper. For that purpose, we allow users to apply for access to our pre-trained ExMed-BERT model on https://doi.org/10.5281/zenodo.7324178 or by sending an email to the corresponding author. Our code is available on https://github.com/SCAI-BIO/ExMed-BERT.

## Data Availability

The data are part of the commercial IBM Explorys Therapeutic dataset. This study's code is accessible to the public at https://github.com/SCAI-BIO/ExMed-BERT.

## Acknowledgments

This research was performed in the context of the ‘COPERIMOplus’ initiative and supported by the Fraunhofer ‘Internal Programs’ under Grant No. Anti-Corona 840266.

*The COPERIMOplus Consortium*: Fraunhofer Data Protection Office (Anne Funck Hansen), Fraunhofer IAIS (Sabine, Kugler Stefan Rüping), Fraunhofer IGD (Jan Burmeister, Jörn Kohlhammer), Fraunhofer IKTS (George Sarau, Silke Christiansen), Fraunhofer IME (Oliver Keminer), Fraunhofer ITMP (Aimo Kannt, Andrea Zaliani, Ann Christina Foldenauer, Carsten Claussen, Eduard Resch, Kevin Frank), Fraunhofer MEVIS (Hendrik Laue, Horst Hahn, Jochen Hirsch, Marco Wischnewski, Matthias Günther, Saulius Archipovas), Fraunhofer SCAI (Alpha Tom Kodamullil, Andre Gemünd, Bruce Schultz, Carina Steinborn, Christian Ebeling, Daniel Domingo Fernández, Helena Hermanowski, Holger Fröhlich, Jürgen Klein, Manuel Lentzen, Marc Jacobs, Martin Hofmann-Apitius, Meike Knieps, Michael Krapp, Philipp Johannes Wendland, Philipp Wegner, Sepehr Golriz Khatami, Stephan Springstubbe, Thomas Linden), ZB MED Information Centre for Life Sciences (Juliane Fluck).

## Contributions

Initiated and supervised project: HF; programmed code and conducted experiments: ML, TL, SV, DK; supervised transfer learning on Austrian hospital data: WL†. Drafted the manuscript: ML, HF. All authors have read and approved the current version of the paper.

## Ethics Declaration

The data analyzed in this study are from two sources. On the one hand, we used the commercial database IBM Explorys Therapeutic Dataset. Since it contains no information about clinical trials and is entirely deidentified, an Institutional Review Board was not required to authorize the use of these data. On the other hand, we utilized data from the Austrian hospitals, which was approved by the Medical University of Graz’s ethics committee (IRB00002556, EK-Number: 30-146 ex 17/18).

## References

[1] Riccardo Miotto et al. “Deep learning for healthcare: review, opportunities and challenges”. eng. In: Briefings in Bioinformatics 19.6 (Nov. 2018), pp. 1236–1246. ISSN: 1477-4054. DOI: 10.1093/bib/bbx044.

[2] Benjamin A. Goldstein et al. “Opportunities and challenges in developing risk prediction models with electronic health records data: a systematic review”. eng. In: Journal of the American Medical Informatics Association: JAMIA 24.1 (Jan. 2017), pp. 198–208. ISSN: 1527-974X. DOI: 10.1093/jamia/ocw042.

[3] Thomas Linden et al. “An Explainable Multimodal Neural Network Architecture for Predicting Epilepsy Comorbidities Based on Administrative Claims Data”. In: Frontiers in Artificial Intelligence 4 (2021). ISSN: 2624-8212. DOI: 10.3389/frai.2021.610197. (Visited on 06/13/2022).

[4] Laila Rasmy et al. CovRNN—A recurrent neural network model for predicting outcomes of COVID-19 patients: model development and validation using EHR data. en. Tech. rep. Type: article. medRxiv, Sept. 2021, p. 2021.09.27.21264121. DOI: 10.1101/2021.09.27.21264121. (Visited on 05/17/2022).

[5] Yikuan Li et al. “BEHRT: Transformer for Electronic Health Records”. en. In: Scientific Reports 10.1 (Apr. 2020). Number: 1 Publisher: Nature Publishing Group, p. 7155. ISSN: 2045-2322. DOI: 10.1038/s41598-020-62922-y. URL: https://www.nature.com/articles/s41598-020-62922-y (visited on 04/29/2022).

[6] Yikuan Li et al. “Hi-BEHRT: Hierarchical Transformer-based model for accurate prediction of clinical events using multimodal longitudinal electronic health records”. In: (2021). Publisher: arXiv Version Number: 1. DOI: 10.48550/ARXIV.2106.11360. 2106.11360.

[7] Laila Rasmy et al. “Med-BERT: pretrained contextualized embeddings on large-scale structured electronic health records for disease prediction”. en. In: npj Digital Medicine 4.1 (May 2021). Number: 1 Publisher: Nature Publishing Group, pp. 1–13. ISSN: 2398-6352. DOI: 10.1038/s41746-021-00455-y. URL: https://www.nature.com/articles/s41746-021-00455-y (visited on 04/29/2022).

[8] Yiwen Meng et al. “Bidirectional Representation Learning From Transformers Using Multimodal Electronic Health Record Data to Predict Depression”. In: IEEE Journal of Biomedical and Health Informatics 25.8 (Aug. 2021). Conference Name: IEEE Journal of Biomedical and Health Informatics, pp. 3121–3129. ISSN: 2168-2208. DOI: 10.1109/JBHI.2021.3063721.

[9] Junyuan Shang et al. Pre-training of Graph Augmented Transformers for Medication Recommendation. 2019.

[10] Emily Alsentzer et al. “Publicly Available Clinical BERT Embeddings”. In: Proceedings of the 2nd Clinical Natural Language Processing Workshop. Minneapolis, Minnesota, USA: Association for Computational Linguistics, June 2019, pp. 72–78. DOI: 10.18653/v1/W19-1909. URL: https://aclanthology.org/W19-1909 (visited on 09/22/2021).

[11] Jinhyuk Lee et al. “BioBERT: a pre-trained biomedical language representation model for biomedical text mining”. In: Bioinformatics (Sept. 2019), btz682. ISSN: 1367-4803, 1460-2059. DOI: 10.1093/bioinformatics/btz682. 1901.08746.

[12] Ahmed Elnaggar et al. ProtTrans: Towards Cracking the Language of Life’s Code Through Self-Supervised Deep Learning and High Performance Computing. May 2021. DOI: 10.48550/arXiv.2007.06225. URL: http://arxiv.org/abs/2007.06225 (visited on 11/25/2022).

[13] Yanrong Ji et al. “DNABERT: pre-trained Bidirectional Encoder Representations from Transformers model for DNA-language in genome”. en. In: Bioinformatics 37.15 (Aug. 2021). Ed. by Janet Kelso, pp. 2112–2120. ISSN: 1367-4803, 1460-2059. DOI: 10.1093/bioinformatics/btab083. URL: https://academic.oup.com/bioinformatics/article/37/15/2112/6128680 (visited on 11/25/2022).

[14] Sumit Madan et al. “Accurate prediction of virus-host protein-protein interactions via a Siamese neural network using deep protein sequence embeddings”. en. In: Patterns 3.9 (Sept. 2022), p. 100551. ISSN: 2666-3899. DOI: 10.1016/j.patter.2022.100551. URL: https://www.sciencedirect.com/science/article/pii/S2666389922001568 (visited on 09/12/2022).

[15] Jacob Devlin et al. “BERT: Pre-training of Deep Bidirectional Transformers for Language Understanding”. In: (May 2019). 1810.04805 [cs].

[16] World Health Organization. ICD-10 : international statistical classification of diseases and related health problems : tenth revision. cs. Tech. rep. ISBN: 9789241546492. World Health Organization, 2004. URL: https://apps.who.int/iris/handle/10665/42980 (visited on 06/07/2022).

[17] Clement J McDonald et al. “LOINC, a Universal Standard for Identifying Laboratory Observations: A 5-Year Update”. en. In: Clinical Chemistry 49.4 (Apr. 2003), pp. 624–633. ISSN: 0009-9147, 1530-8561. DOI: 10.1373/49.4.624. URL: https://academic.oup.com/clinchem/article/49/4/624/5641953 (visited on 11/22/2022).

[18] The Central Role of the Propensity Score in Observational Studies for Causal Effects. URL: https://dash.harvard.edu/handle/1/3382855 (visited on 09/21/2022).

[19] Paul R. Rosenbaum. “Model-Based Direct Adjustment”. In: Journal of the American Statistical Association 82.398 (June 1987), pp. 387–394. ISSN: 0162-1459. DOI: 10.1080/01621459.1987.10478441. (Visited on 09/21/2022).

[20] Peter C. Austin and Muhammad M. Mamdani. “A comparison of propensity score methods: a case-study estimating the effectiveness of post-AMI statin use”. en. In: Statistics in Medicine 25.12 (2006), pp. 2084–2106. ISSN: 1097-0258. DOI: 10.1002/sim.2328. (Visited on 09/21/2022).

[21] Adrienne Kline. psmpy: Propensity score matching for python and graphical plots.

[22] Stuart J Nelson et al. “Normalized names for clinical drugs: RxNorm at 6 years”. en. In: Journal of the American Medical Informatics Association 18.4 (July 2011), pp. 441–448. ISSN: 1067-5027, 1527-974X. DOI: 10.1136/amiajnl-2011-000116. (Visited on 11/15/2022).

[23] Patrick Wu et al. Developing and Evaluating Mappings of ICD-10 and ICD-10-CM codes to Phecodes. en. Tech. rep. Section: New Results Type: article. bioRxiv, Nov. 2018, p. 462077. DOI: 10.1101/462077. (Visited on 06/07/2022).

[24] WHO Collaborating Centre for Drug Statistics Methodology, ATC classification index with DDDs. Oslo, Norway 2021. 2022. URL: https://www.whocc.no/atc_ddd_index_and_guidelines/atc_ddd_index/ (visited on 06/08/2022).

[25] Takuya Akiba et al. “Optuna: A Next-generation Hyperparameter Optimization Framework”. In: Proceedings of the 25rd ACM SIGKDD International Conference on Knowledge Discovery and Data Mining. 2019.

[26] Missing Data Imputation for Python - missingpy. Publisher: Ashim Bhattarai. URL: https://pypi.org /project/missingpy/ (visited on 06/08/2022).

[27] Mukund Sundararajan, Ankur Taly, and Qiqi Yan. Axiomatic Attribution for Deep Networks. Number: arXiv:1703.01365 arXiv:1703.01365 [cs]. June 2017. DOI: 10.48550/arXiv.1703.01365. 1703.01 365.

[28] Marco Scutari. “Learning Bayesian Networks with the bnlearn R Package”. In: Journal of Statistical Software 35.3 (2010), pp. 1–22. DOI: 10.18637/jss.v035.i03.

[29] Fred Glover and Claude McMillan. “The general employee scheduling problem. An integration of MS and AI”. en. In: Computers & Operations Research. Applications of Integer Programming 13.5 (Jan. 1986), pp. 563–573. ISSN: 0305-0548. DOI: 10.1016/0305-0548(86)90050-X. URL: https://www.sciencedirect.com/science/article/pii/030505488690050X (visited on 09/27/2022).

[30] Tabu Search—Part I | ORSA Journal on Computing. DOI: 10.1287/ijoc.1.3.190. (Visited on 09/27/2022).

[31] Z. H. Israili. “Clinical pharmacokinetics of angiotensin II (AT1) receptor blockers in hypertension”. en. In: Journal of Human Hypertension 14.1 (Apr. 2000). Number: 1 Publisher: Nature Publishing Group, S73–S86. ISSN: 1476-5527. DOI: 10.1038/sj.jhh.1000991. URL: https://www.nature.com/articles/1000991 (visited on 11/09/2022).

[32] Riccardo Vigneri and Ira D Goldfine. “Role of metformin in treatment of diabetes mellitus”. en. In: Diabetes Care 10.1 (Jan. 1987), pp. 118–122. ISSN: 0149-5992, 1935-5548. DOI: 10.2337/diacare.10.1.118. URL: https://diabetesjournals.org/care/article/10/1/118/790/Role-of-metformi n-in-treatment-of-diabetes (visited on 11/09/2022).

[33] Wei-jie Guan et al. “Comorbidity and its impact on 1590 patients with COVID-19 in China: a nationwide analysis”. en. In: European Respiratory Journal 55.5 (May 2020). Publisher: European Respiratory Society Section: Original Articles. ISSN: 0903-1936, 1399-3003. DOI: 10.1183/13993003.00547-2020. URL: https://erj.ersjournals.com/content/55/5/2000547 (visited on 06/07/2022).

[34] Ana C. Tahira, Sergio Verjovski-Almeida, and Sergio T. Ferreira. “Dementia is an age-independent risk factor for severity and death in COVID-19 inpatients”. en. In: Alzheimer’s & Dementia 17.11 (2021), pp. 1818–1831. ISSN: 1552-5279. DOI: 10.1002/alz.12352. (Visited on 09/21/2022).

[35] Sofia Toniolo et al. “Dementia and COVID-19, a Bidirectional Liaison: Risk Factors, Biomarkers, and Optimal Health Care”. en. In: Journal of Alzheimer’s Disease 82.3 (Jan. 2021). Publisher: IOS Press, pp. 883–898. ISSN: 1387-2877. DOI: 10.3233/JAD-210335. URL: https://content.iospress.com/articles /journal-of-alzheimers-disease/jad210335 (visited on 06/07/2022).

[36] Slobodan Peric and Thomas M. Stulnig. “Diabetes and COVID-19”. en. In: Wiener klinische Wochenschrift 132.13 (July 2020), pp. 356–361. ISSN: 1613-7671. DOI: 10.1007/s00508-020-01672-3. (Visited on 06/08/2022).

[37] Fien Demeulemeester et al. “Obesity as a Risk Factor for Severe COVID-19 and Complications: A Review”. en. In: Cells 10.4 (Apr. 2021). Number: 4 Publisher: Multidisciplinary Digital Publishing Institute, p. 933. ISSN: 2073-4409. DOI: 10.3390/cells10040933. URL: https://www.mdpi.com/2073-4409/10/4/933 (visited on 09/21/2022).

[38] See Kwok et al. “Obesity: A critical risk factor in the COVID-19 pandemic”. en. In: Clinical Obesity 10.6 (2020), e12403. ISSN: 1758-8111. DOI: 10.1111/cob.12403. (Visited on 09/21/2022).

[39] Rachel Concha, Elan Ohayon, and Ann Lam. “Neuroinflammation in COVID-19 and ADRD: Similarities, differences, and interactions”. en. In: Alzheimer’s & Dementia 17.S3 (2021), e056282. ISSN: 1552-5279. DOI: 10.1002/alz.056282. (Visited on 09/27/2022).

[40] Charlotte Steenblock et al. “COVID-19 and metabolic disease: mechanisms and clinical management”. eng. In: The Lancet. Diabetes & Endocrinology 9.11 (Nov. 2021), pp. 786–798. ISSN: 2213-8595. DOI: 10.1016/S2213-8587(21)00244-8.

[41] Nicola Lazzarini et al. “A machine learning model on Real World Data for predicting progression to Acute Respiratory Distress Syndrome (ARDS) among COVID-19 patients”. eng. In: PloS One 17.7 (2022), e0271227. ISSN: 1932-6203. DOI: 10.1371/journal.pone.0271227.

[42] Itai Bendavid et al. “A novel machine learning model to predict respiratory failure and invasive mechanical ventilation in critically ill patients suffering from COVID-19”. en. In: Scientific Reports 12.1 (June 2022). Number: 1 Publisher: Nature Publishing Group, p. 10573. ISSN: 2045-2322. DOI: 10.1038/s41598-022-14758-x. URL: https://www.nature.com/articles/s41598-022-14758-x (visited on 11/21/2022).

[43] Lakshya Singhal et al. “eARDS: A multi-center validation of an interpretable machine learning algorithm of early onset Acute Respiratory Distress Syndrome (ARDS) among critically ill adults with COVID-19”. en. In: PLOS ONE 16.9 (Sept. 2021). Publisher: Public Library of Science, e0257056. ISSN: 1932-6203. DOI: 10.1371/journal.pone.0257056. URL: https://journals.plos.org/plosone/article?id=10.1371/journal.pone.0257056 (visited on 09/20/2022).

[44] Yingzhen Du et al. “Clinical Features of 85 Fatal Cases of COVID-19 from Wuhan. A Retrospective Observational Study”. In: American Journal of Respiratory and Critical Care Medicine 201.11 (June 2020). Publisher: American Thoracic Society - AJRCCM, pp. 1372–1379. ISSN: 1073-449X. DOI: 10.1164/rccm.202003-0543OC. (Visited on 06/07/2022).

[45] Bo Li et al. “Prevalence and impact of cardiovascular metabolic diseases on COVID-19 in China”. en. In: Clinical Research in Cardiology 109.5 (May 2020), pp. 531–538. ISSN: 1861-0692. DOI: 10.1007/s00392-020-01626-9. (Visited on 06/07/2022).

[46] Thomas Linden et al. “Machine Learning Based Prediction of COVID-19 Mortality Suggests Repositioning of Anticancer Drug for Treating Severe Cases”. en. In: Artificial Intelligence in the Life Sciences 1 (Dec. 2021), p. 100020. ISSN: 2667-3185. DOI: 10.1016/j.ailsci.2021.100020. URL: https://www.sciencedirect.com/science/article/pii/S2667318521000209 (visited on 11/22/2022).

