## Supplementary Material for "A Transformer-Based Model Trained on Large Scale Claims Data for Prediction of Severe COVID-19 Disease Progression"

### A. Data

#### A. Criteria for COVID confirmation

- U07.01 code as a diagnosis in medical history
- Positive test results for one of the following tests (LOINC-encoded)
  - 94500-6
  - 41001-9
  - 82162-9
  - 82161-1
  - 82164-5
  - 82163-7
  - 62423-9
  - 94307-6
  - 94308-4
  - 94309-2
  - 94310-0
  - 94311-8
  - 94312-6
  - 94313-4
  - 94314-2
  - 94315-9
  - 94316-7
  - 94500-6
  - 94502-2
  - 94505-5
  - 94506-3
  - 94507-1
  - 94508-9
  - 94509-7
  - 94510-5
  - 94511-3
  - 94532-9
  - 94533-7
  - 94534-5
  - 94547-7

### B. Filtering strategy

Supplementary Figure A.1. Filtering strategy. Shown are the different subsets of the IBM Explorys Therapeutic dataset, which were used for the experiments of this study.

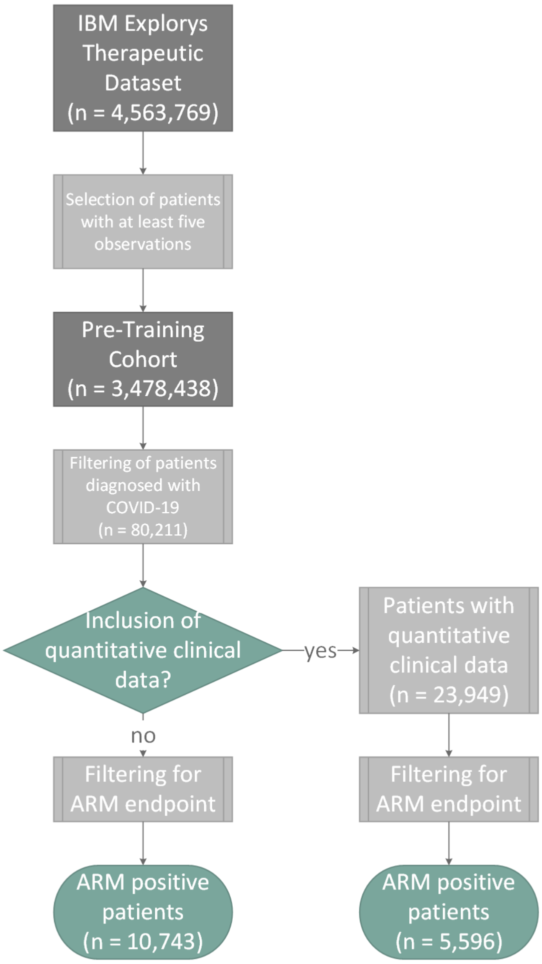

### B. Experiments

#### A. Tuned Hyperparameters

| | Hyperparameter | Range |
|---|---|---|
| ExMed-BERT | Learning rate | [1e-5, 5e-3] |
| | Batch size | 8, 16, 32 (categorical) |
| | Warmup ratio | 0,0.05, 0.1 (categorical) |
| | Weight decay | [1e-5, 1e-1] |
| | (Number of RNN layers) | [1, 4] |
| XGBoost | Maximum depth | [4, 8] |
| | Learning rate | [1e-5, 1] |
| | Subsample | [0.5, 1] |
| | Rate drop | [0, 0.2] |
| | Scale positive weight | [1, 14] |
| Random Forest | Number of estimators | [50, 1000] |
| | Maximum depth | [2, 200] |
| | Minimum samples per split | [2, 150] |
| | Minimum samples per leaf | [1, 60] |
| | Maximum features | sqrt, log (categorical) |

Supplementary Table B.1. Tuned Hyperparameters. Shown are all parameters that were tuned during hyperparameter optimization.

#### B. Pre-training of ExMed-BERT

Supplementary Figure B.1. Pre-training Metrics. Shown are the masked language modeling (MLM) accuracy and the F1 score to predict a prolonged length of stay (PLOS) in a hospital. We trained our model for 4.5 Million steps before we modified the model's vocabulary (addition of 61 ATC codes) and trained the model for further steps.

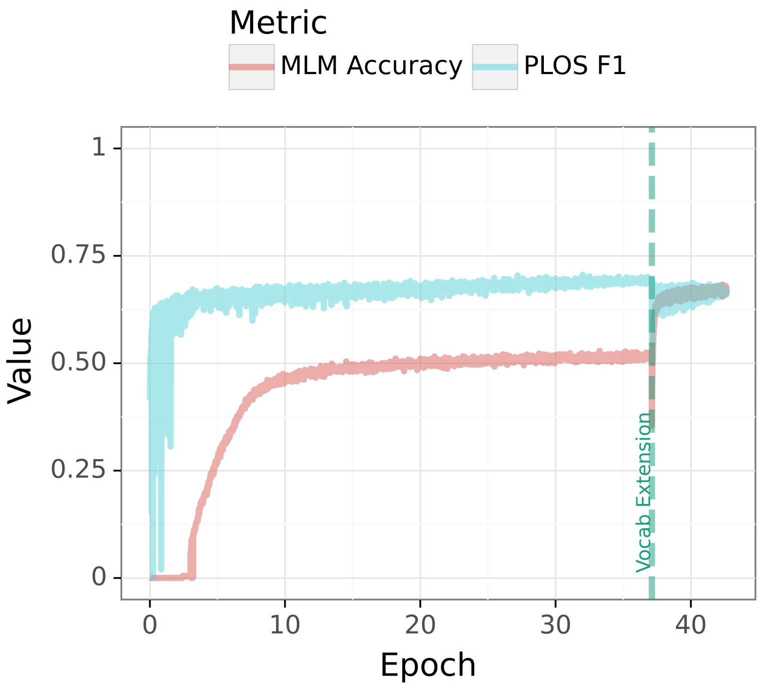

#### C. Adaption and Evaluation of Risk Models on data from the Austrian Hospital Group KAGes

In addition to the IBM Explorys Therapeutic dataset, we used data from the Austrian hospital group KAGes to assess the transfer-learning capabilities of our ExMed-BERT model. As described in the methods section, we processed the data from the 6335 inpatients as we did the IBM Explorys data. We mapped ICD9/10 encoded diagnoses to Phecodes and used the fourth ATC level for drugs. The models fine-tuned on IBM data were then used in a transfer-learning approach in which we continued training on KAGes data. Due to computational constraints, we used the best hyperparameters found on the IBM Explorys data. To account for the limited amount of available data, we used 5-fold cross-validation. In addition to the ExMed-BERT model, we trained a new RF model as a comparison. The results in Supplementary Table B.2 show that there is no significant difference between the RF and ExMed-BERT models. In all cases, the AUROC is approximately 60 %, and the AUPR is 9 %.

Supplementary Table B.2. Results of the fine-tuned models for the KAGes data. Areas under the Receiver Operator Characteristic Curve (AUROC) and the Precision-Recall Curve (AUPR). The best results per column are highlighted in bold.

| | AUROC | AUPR |
|---|---|---|
| RF | 60.0 ± 1.8 | 9.0 ± 1.2 |
| ExMed-BERT-FFN | 60.1 ± 1.7 | 9.0 ± 0.7 |
| ExMed-BERT-GRU | 59.0 ± 3.5 | 9.0 ± 0.9 |
| ExMed-BERT-LSTM | 60.0 ± 3.0 | 9.0 ± 0.8 |
